# Duration of Protection Against SARS-CoV-2 Reinfection and Associated Risk of Reinfection Assessed with Real-World Data

**DOI:** 10.1101/2022.02.25.22271515

**Authors:** Shannon L. Reynolds, Harvey W. Kaufman, William A. Meyer, Chris Bush, Oren Cohen, Kathy Cronin, Carly Kabelac, Sandy Leonard, Steve Anderson, Valentina Petkov, Douglas Lowy, Norman Sharpless, Lynne Penberthy

## Abstract

**Importance:** Better understanding of the protective duration of prior SARS-CoV-2 infection against reinfection is needed.

**Objective:** Primary: To assess the durability of immunity to SARS-CoV-2 reinfection among initially unvaccinated individuals with previous SARS-CoV-2 infection. Secondary: Evaluate the crude SARS-CoV-2 reinfection rate and associated characteristics.

**Design and Setting:** Retrospective observational study of HealthVerity data among 144,678,382 individuals, during the pandemic era through April 2021.

**Participants:** Individuals studied had SARS-CoV-2 molecular diagnostic or antibody index test results from February 29 through December 9, 2020, with ≥365 days of pre-index continuous closed medical enrollment, claims, or electronic health record activity.

**Main Outcome(s) and Measure(s):** Rates of reinfection among index-positive individuals were compared to rates of infection among index-negative individuals. Factors associated with reinfection were evaluated using multivariable logistic regression. For both objectives, the outcome was a subsequent positive molecular diagnostic test result.

**Results:** Among 22,786,982 individuals with index SARS-CoV-2 laboratory test data (2,023,341 index positive), the crude rate of reinfection during follow-up was significantly lower (9.89/1,000-person years) than that of primary infection (78.39/1,000 person years). Consistent with prior findings, the risk of reinfection among index-positive individuals was 87% lower than the risk of infection among index-negative individuals (hazard ratio, 0.13; 95% CI, 0.13, 0.13). The cumulative incidence of reinfection among index-positive individuals and infection among index-negative individuals was 0.85% (95% CI: 0.82%, 0.88%) and 6.2% (95% CI: 6.1%, 6.3%), respectively, over follow-up of 375 days. The duration of protection against reinfection was stable over the median 5 months and up to 1-year follow-up interval. Factors associated with an increased reinfection risk included older age, comorbid immunologic conditions, and living in congregate care settings; healthcare workers had a decreased reinfection risk.

**Conclusions and Relevance:** This large US population-based study demonstrates that SARS-CoV-2 reinfection is uncommon among individuals with laboratory evidence of a previous infection. Protection from SARS-CoV-2 reinfection is stable up to one year. Reinfection risk was primarily associated with age 85+ years, comorbid immunologic conditions and living in congregate care settings; healthcare workers demonstrated a decreased reinfection risk. These findings suggest that infection induced immunity is durable for variants circulating prior to Delta.

**Key Points:** *Question:* How long does prior SARS-CoV-2 infection provide protection against SARS-CoV-2 reinfection?

*Finding:* Among >22 million individuals tested February 2020 through April 2021, the relative risk of reinfection among those with prior infection was 87% lower than the risk of infection among individuals without prior infection. This protection was durable for up to a year. Factors associated with increased likelihood of reinfection included older age (85+ years), comorbid immunologic conditions, and living in congregate care settings; healthcare workers had lower risk.

*Meaning:* Prior SARS-CoV-2 infection provides a durable, high relative degree of protection against reinfection.

## Rationale and Background

To date, over 329 million confirmed cases of COVID-19 have been diagnosed globally.^1^ Survivors have a risk of reinfection that can be associated with serious clinical outcomes.^2, 3^ Our previous study of a national U.S. cohort found that seropositivity was associated with reduced risk of subsequent infection over a relatively short interval.^4^ That study demonstrated that at >90 days after an index SARS-CoV-2 antibody test, the ratio of positive nucleic acid amplification test (NAAT) results between those who were index SARS-CoV-2 positive versus index negative individuals was 0.1, suggesting prior infection provided ~90% protection from reinfection. Similar results were observed using real-world data (RWD) and different study designs. ^5–9^ Research has also demonstrated that serum SARS-CoV-2 neutralizing antibodies and T cell immunity correlate with protection against infection and reinfection.^10, 11^

Previous short-term studies relying on RWD indicate prior SARS-CoV-2 infection may offer protection for at least 7 months.^5–9^ Existing studies of reinfection have limited follow-up time for observing reinfection and often limited study population size and geography. For instance, many are limited to healthcare workers or students and lack generalizability to other at-risk populations, such as the elderly and individuals with comorbidities.^5–9^ Accordingly, it is unclear whether the duration of protection associated with SARS-CoV-2 seropositivity may be substantially longer, and the impact of patient-level characteristics on the risk of reinfection is also unknown.

Large-scale RWD offer an opportunity to study patterns of infection and reinfection, available longitudinally at the individual level, making it possible to study the experiences of a seropositive population with COVID-19 in near-real time. Further advantages of real-world database studies include maximizing sample size, the ability to evaluate risk within subgroups and specific patient characteristics, and greater data capture of the patient experience over time. The primary objective of this study was to estimate the duration of protection provided against laboratory confirmed SARS-CoV-2 reinfection among index SARS-CoV-2 positive individuals compared to the risk of infection among index SARS-CoV-2 negative individuals. The secondary objective was to estimate the rate of reinfection among those previously identified as SARS-CoV-2 positive and evaluate demographic and comorbid characteristics associated with the risk of SARS-CoV-2 reinfection.

## Methods

### Data sources

The study population began with 144,678,382 individuals derived from US RWD sources curated by HealthVerity and with records of medical services obtained from February 29, 2020, through April 30, 2021. This dataset aggregates multiple unique record sources across commercial laboratory databases (including an estimated 60% of aggregate SARS-CoV-2 testing performed in the US), medical claims (both open and closed claims), pharmacy databases (both claims and retail), hospital chargemaster (CDM) and outpatient electronic health records (EHR). Patient records, using a unique, interoperable, de-identified token, are linked across laboratory test results, medical and pharmacy claims, CDM, and any available EHR. The data included in this study are compliant with HIPAA such that no patient can be reidentified. The study was deemed exempt by the New England Institutional Review Board (#1-9757-1).

### Study Design

#### Study Population

The dataset included records for individuals with an index NAAT or SARS-CoV-2 antibody test from February 29, 2020 (when SARS-CoV-2 NAAT first became available for molecular diagnostic testing), through December 9, 2020 (prior to COVID-19 vaccine introduction). Individuals included based on an antibody test result could enter starting on April 15, 2020, the earliest availability of an antibody test with US Food and Drug Administration (FDA) Emergency Use Authorization (EUA).

For the primary study objective, a cohort of SARS-CoV-2-tested individuals was identified. Individuals entered based on their first SARS-CoV-2 antibody or NAAT test result (index date). Individuals were required to have ≥12 months of pre-index continuous closed medical enrollment, claims or electronic health record (EHR) activity. Individuals with discordant test results on the same day were excluded. Additionally, those who tested SARS-CoV-2 NAAT-positive within 60 days following their initial test results (applied only to the index negative group) or were lost to follow-up during those first 60 days were excluded.

For the secondary objective, a cohort of individuals with any record of SARS-CoV-2-positive test results were identified. Individuals entered based on their first SARS-CoV-2 antibody or NAAT-positive test result from February 29, 2020, through December 9, 2020. Individuals were required to have ≥12 months of pre-index continuous closed medical enrollment or activity as described above. Individuals with discordant test results on the same day were excluded.

For both objectives, the baseline period was the 12 months prior to the index test date. All available data in this period were used to identify baseline characteristics and comorbid conditions.

#### Exposure, Outcome and Covariates

For the primary objective, individuals entered the cohort upon their first SARS-CoV-2 NAAT or antibody test; those with a positive result were classified as the index-positive group (i.e., established SARS-CoV-2 infection) while individuals whose first NAAT or antibody test result was negative were the index-negative group (i.e., without laboratory evidence of SARS-CoV-2 infection). For the secondary objective, the cohort consisted only of individuals who had a SARS-CoV-2 positive test result.

The outcome of interest for both objectives was a SARS-CoV-2 positive NAAT result occurring at >60 days after the index test result date. The follow-up time interval began on the 61st day post index date, rather than the patient index date, to avoid misclassifying individuals who experienced prolonged viral RNA shedding in the weeks after their initial infection as having the outcome.^4, 10, 11^ Individuals were followed until the outcome of interest or the earliest occurrence of inpatient death, outcome of interest, end of available data, end of their medical plan enrollment or end of activity.

#### Sensitivity analysis

To demonstrate the robustness of our laboratory confirmed outcome definition, we examined International Classification of Diseases (ICD)-10 diagnosis codes U07.1 or U07.2 in addition to the laboratory test results for outcome ascertainment as ICD codes are commonly used in real-world research to define occurrence of COVID-19 and may affect the outcome rate.^12^

#### Statistical Analysis

Variables included as baseline characteristics and model covariates are defined in the supplement and were reported descriptively (eMethods1 and eMethods2). Continuous variables are presented as means (with standard deviation) and/or medians (with interquartile range). Categorical variables are presented as absolute and relative frequencies. Demographic covariates were assessed on the index date. Regions are defined by the US Census Bureau.^13^ Presence of comorbidities was assessed during the 12-month baseline period prior to the index date, unless specified otherwise in the supplement. For the primary objective, missing data that occurred in covariates or descriptive variables were classified using a missing data indicator.

For the primary objective, propensity score matching, estimated via logistic regression, was used to adjust for potential confounding between index-positive and index-negative individuals on the index date. Individuals were matched by propensity score on a 1:1 basis using a caliper of 1.0%. Variables included in the propensity-score model are defined in the supplement (eMethods 1). Covariate balance post-matching was evaluated using standardized differences with a threshold of <0.10 to indicate well-balanced differences.^14^

For the primary objective, we report the crude rate of SARS-CoV-2 infection per 1,000 person-years (PY) and present cumulative incidence curves. We used Cox proportional-hazard regression to estimate the hazard ratios (HR) and 95% CI of the primary outcome for index-positive versus index-negative individuals during follow-up.

For the secondary objective, we estimated the crude rate of reinfection per 1,000 PY in the overall population and by demographic subgroups. Association between individual risk factors and reinfection was explored using a Cox proportional hazard regression model. Characteristics of interest included individual demographics and comorbid conditions. Missing data elements that occurred in characteristics of interest were classified as the most frequently occurring value for categorical variables. We prespecified a 2-tailed alpha of 0.05 to establish statistical significance. Data were analyzed using the previously validated Aetion Evidence Platform version r4.27.0.20210609 and R version 3.4.2.

## Results

For the primary objective, 27,070,023 individuals met the inclusion criteria, of which 7,501 died and 4,275,540 disenrolled between index and start of follow-up; 22,786,982 individuals started follow-up 61 days post-index date. Of these, 2,023,341 (9%) were SARS-CoV-2 index-positive and 20,763,641 (91%) were SARS-CoV-2 index-negative. There were similar age and sex distributions between index-positive and index-negative individuals: a mean age of 43 and 45 years with 57% and 60% female, respectively. The index-negative population exhibited slightly worse comorbid illness (average Charlson-Quan score of 0.55 versus 0.49 for the index-positive). Median follow-up time was 149 days and 162 days for index-positive and index-negative groups, respectively. Index-positive individuals had fewer NAAT tests during follow-up than index-negative individuals (mean (SD) 0.40 (2.55) vs. 0.78 (3.29) respectively). All index-positive individuals were matched to an index-negative individual in the propensity-score model, with 2,023,341 individuals in each group. Characteristics were well balanced before and after matching (Table 1).

**Table 1.**
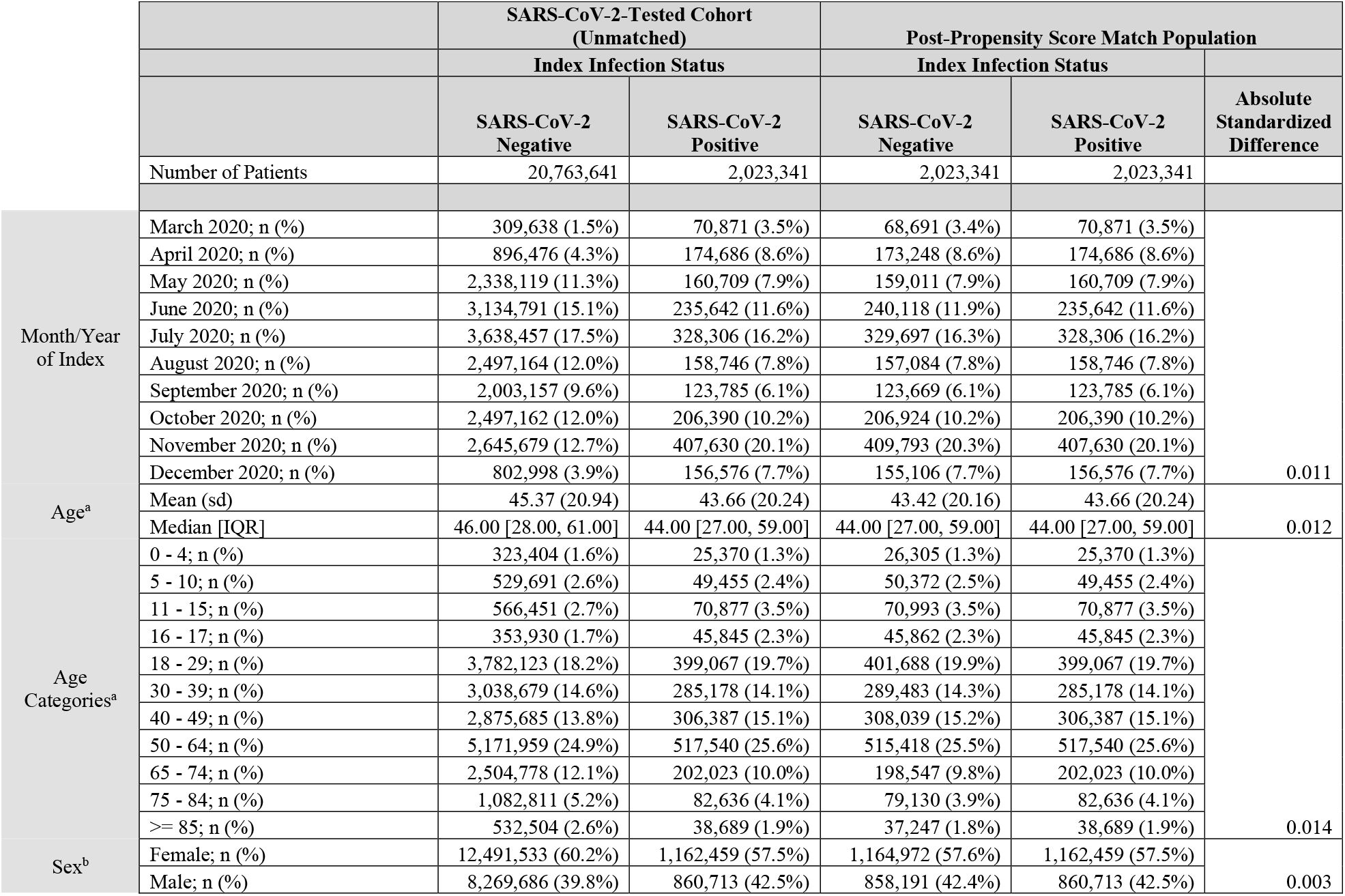
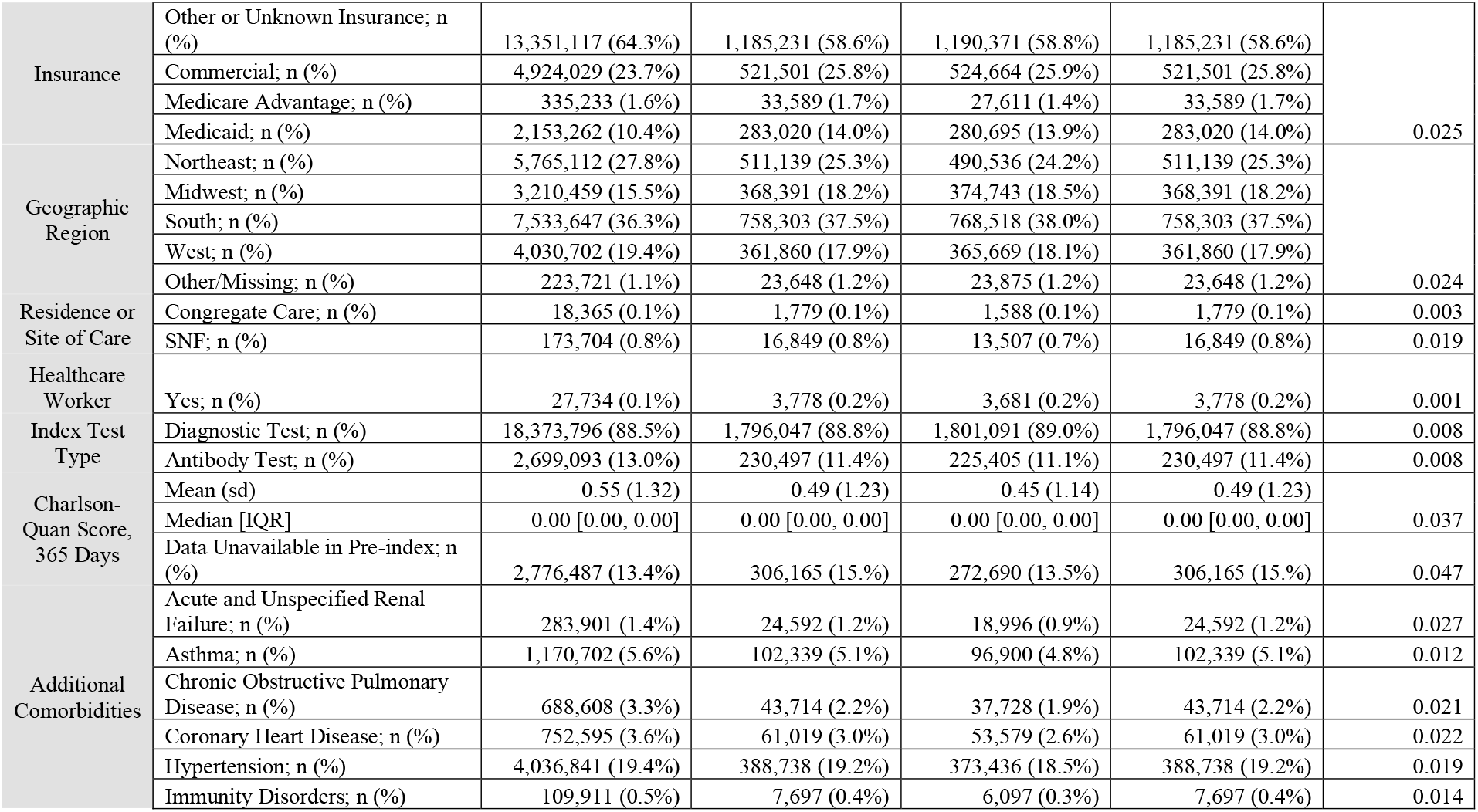
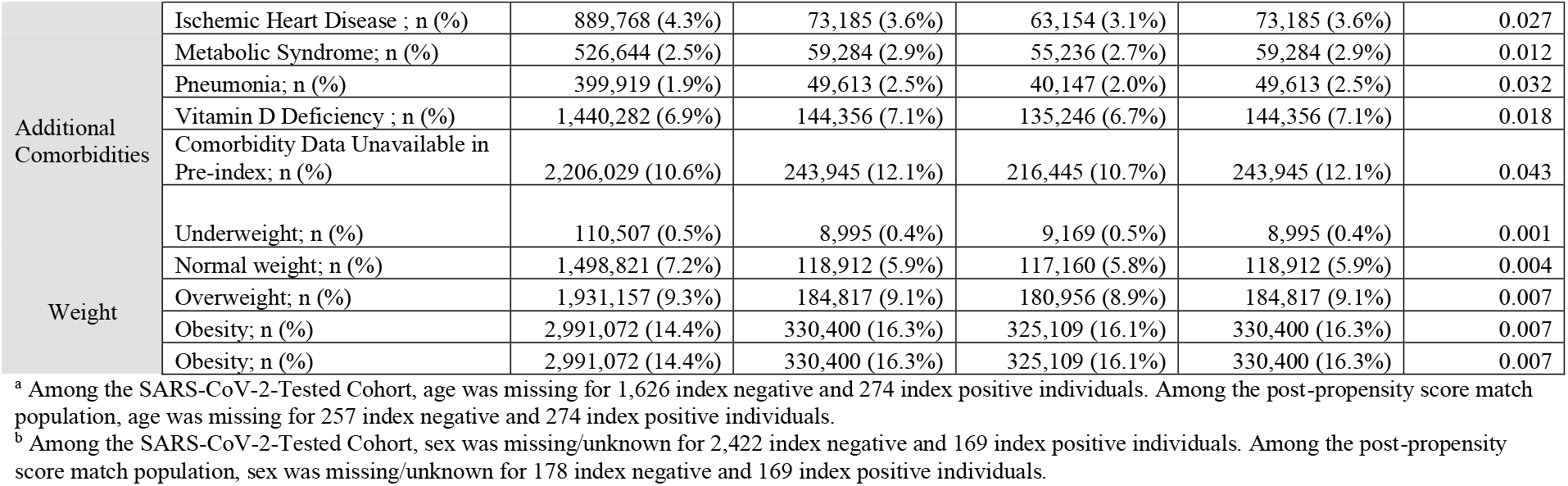
Demographics and baseline comorbidities in the 365 days prior to first SARS-CoV-2 test among a SARS-CoV-2-tested cohort and among a post-propensity score match population identified between February 29th, 2020 through December 9th, 2020

A total of 737,742 cases of infection were observed in the unmatched population over the follow-up period; 8,869 (1.2%) occurred in the index-positive group. The crude rate of reinfection over follow-up was substantially lower in the index-positive group (9.89 per 1,000-person year) than in the index-negative group (78.39 per 1,000 PY) (Table 2). The cumulative incidence of reinfection was 0.85% (95% CI: 0.82%, 0.88%) among index-positive individuals and for infection the rate was 6.2% (95% CI: 6.1%, 6.3%) among index-negative individuals over a follow-up of 375 days from the index date (Figure 1). The risk of reinfection was stable over the median 5 months of follow-up and up to one year among index-positive individuals (n=1,821,183 at 4 months, n=859,824 at 8 months and n=147,458 at 12 months). After adjustment for baseline demographic and comorbid characteristics, the risk of reinfection in the index-positive group was 87% lower than the risk of infection in the index-negative group (HR= 0.13, 95% CI: (0.13, 0.13)). A propensity-score matched analysis provided a nearly identical estimate of the degree and duration of protection. The adjusted risk of infection over follow-up time was relatively stable (6-8 months post-index (HR=0.12, 95% CI: (0.12, 0.13); 8-10 months post-index (HR=0.17, 95% CI: 0.15, 0.18); 10–12 months post-index (HR=0.15, 95% CI: 0.13, 0.18)). Stratified by index-test type (antibody only (n=2,617,139), NAAT only (n=19,857,392) or both (n=312,451), the adjusted risk of reinfection was similar to the pooled estimate (HR=0.14, 95% CI: (0.13, 0.15) for antibody only; HR=0.13 95% CI: (0.13, 0.13) for NAAT only; HR=0.13, 95% CI: (0.08, 0.20) for both on index). These data suggest that SARS-CoV-2 infection provides substantial protection from reinfection for at least 5 months and up to one year from recovery.

**Table 2.**
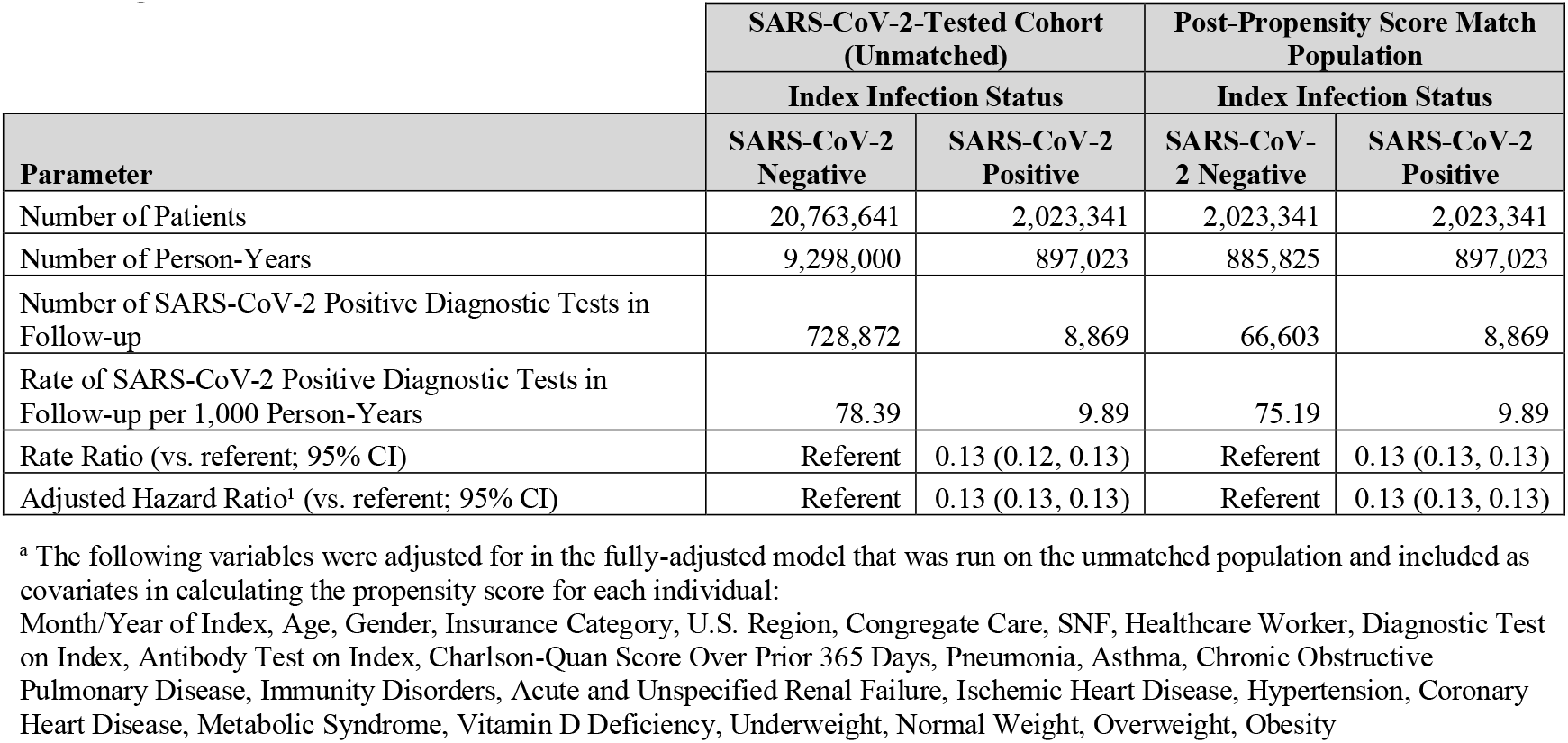
Rate of SARS-CoV-2 positive diagnostic tests in follow-up (April 30th, 2020 through April 30th, 2021) stratified by index SARS-CoV-2 infection status with rate ratio and hazard ratio estimates among SARS-CoV-2 positive individuals vs. SARS CoV-2 negative individuals

| Parameter | SARS-CoV-2-Tested Cohort<br>(Unmatched) |  | Post-Propensity Score Match<br>Population |  |
| --- | --- | --- | --- | --- |
|  | Index Infection Status |  | Index Infection Status |  |
|  | SARS-CoV-2<br>Negative | SARS-CoV-2<br>Positive | SARS-CoV-2<br>Negative | SARS-CoV-2<br>Positive |
| Number of Patients | 20,763,641 | 2,023,341 | 2,023,341 | 2,023,341 |
| Number of Person-Years | 9,298,000 | 897,023 | 885,825 | 897,023 |
| Number of SARS-CoV-2 Positive Diagnostic Tests in<br>Follow-up | 728,872 | 8,869 | 66,603 | 8,869 |
| Rate of SARS-CoV-2 Positive Diagnostic Tests in<br>Follow-up per 1,000 Person-Years | 78.39 | 9.89 | 75.19 | 9.89 |
| Rate Ratio (vs. referent; 95% CI) | Referent | 0.13 (0.12, 0.13) | Referent | 0.13 (0.13, 0.13) |
| Adjusted Hazard Ratio<sup>a</sup> (vs. referent; 95% CI) | Referent | 0.13 (0.13, 0.13) | Referent | 0.13 (0.13, 0.13) |

<sup>a</sup> The following variables were adjusted for in the fully-adjusted model that was run on the unmatched population and included as covariates in calculating the propensity score for each individual:
Month/Year of Index, Age, Gender, Insurance Category, U.S. Region, Congregate Care, SNF, Healthcare Worker, Diagnostic Test on Index, Antibody Test on Index, Charlson-Quan Score Over Prior 365 Days, Pneumonia, Asthma, Chronic Obstructive Pulmonary Disease, Immunity Disorders, Acute and Unspecified Renal Failure, Ischemic Heart Disease, Hypertension, Coronary Heart Disease, Metabolic Syndrome, Vitamin D Deficiency, Underweight, Normal Weight, Overweight, Obesity

**Figure 1.**
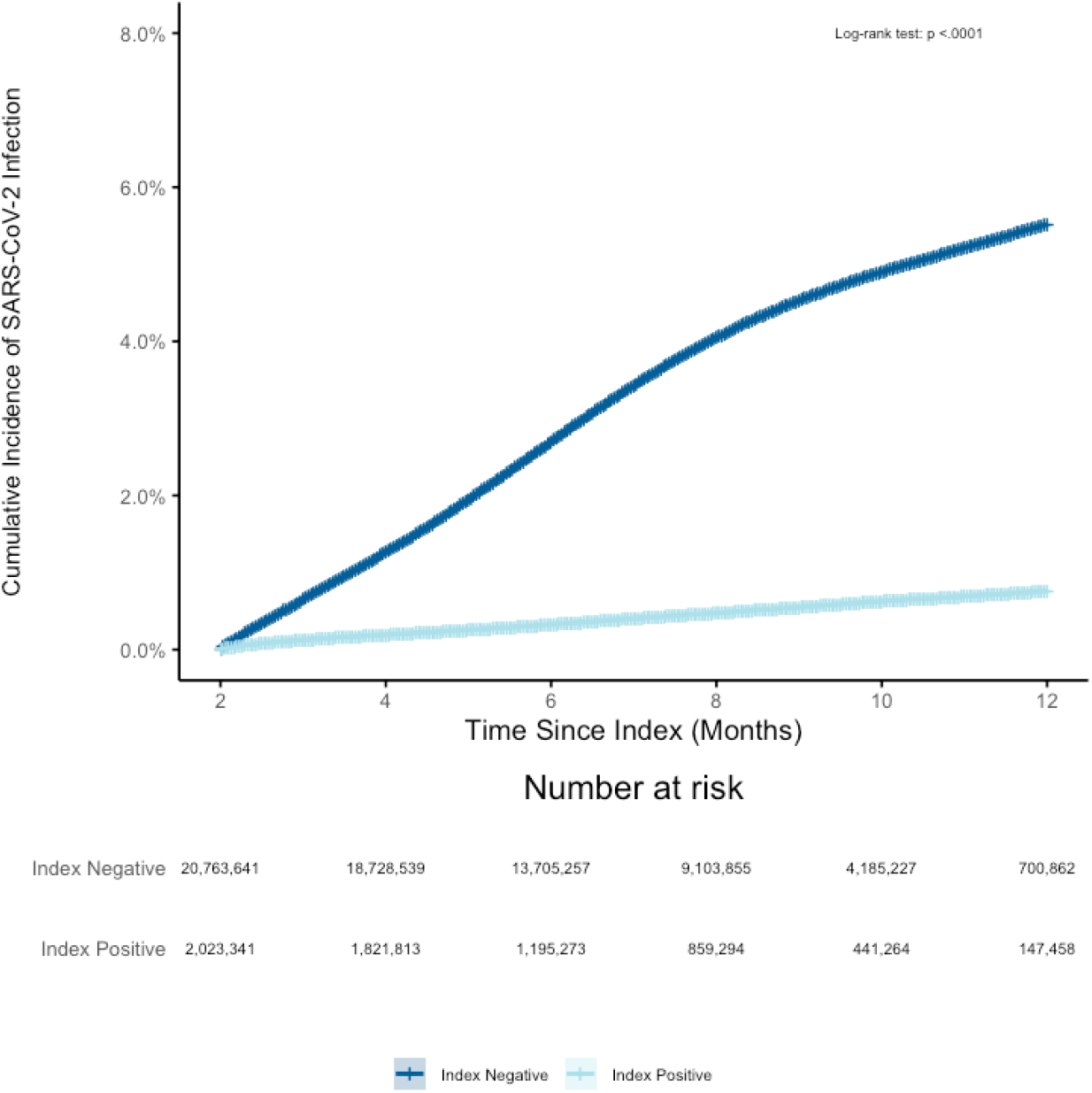
Cumulative incidence of SARS-CoV-2 infection from April 30th, 2020 through April 30th, 2021 among SARS-CoV-2 index positive vs SARS-CoV-2 index negative individuals. There were 71,921 individuals who indexed negative but had a subsequent positive test result within the following 60 days. Of the 27,070,023 index positive and negative individuals, within the first 60 days from index date, 7,501 died in an inpatient setting and 4,275,540 were disenrolled or had no additional claims or EHR activity resulting in 22,786,982 who started follow-up on day 61.

To demonstrate the robustness of laboratory-confirmed COVID defining our outcome, a sensitivity analysis incorporating ICD-10 diagnostic codes to the primary outcome was performed in the primary analysis cohort. The direction of the association was consistent with the primary analysis; however, we estimated only a 27% reduction (fully-adjusted HR= 0.73 (95% CI: 0.72, 0.73)) in risk of reinfection among index-positive individuals versus infection among index-negative individuals, compared to an 87% reduction in the main analysis.

To test the robustness of the prolonged viral shedding period definition on outcome estimates and to align to other previously published literature, we examined a 90-day exclusion period in addition to the primary definition of 60 days.^15–17^ Findings (not shown) were nearly identical to the primary definition results indicating minimal bias in 60 versus 90-day viral shedding exclusion period definitions.

For the secondary study objective, 3,213,214 individuals met the criteria for the SARS-CoV-2-positive cohort, of whom 2,535,887 individuals were not censored due to death, disenrollment, or the end of data prior to beginning follow-up 61 days after their index date. The mean age was 44 years, 58.1% were female and the average Charlson-Quan score was 0.51 (Table 3). Median follow-up time was 155 days. Characteristics were similar between patients who were reinfected and those who were not.

**Table 3.**
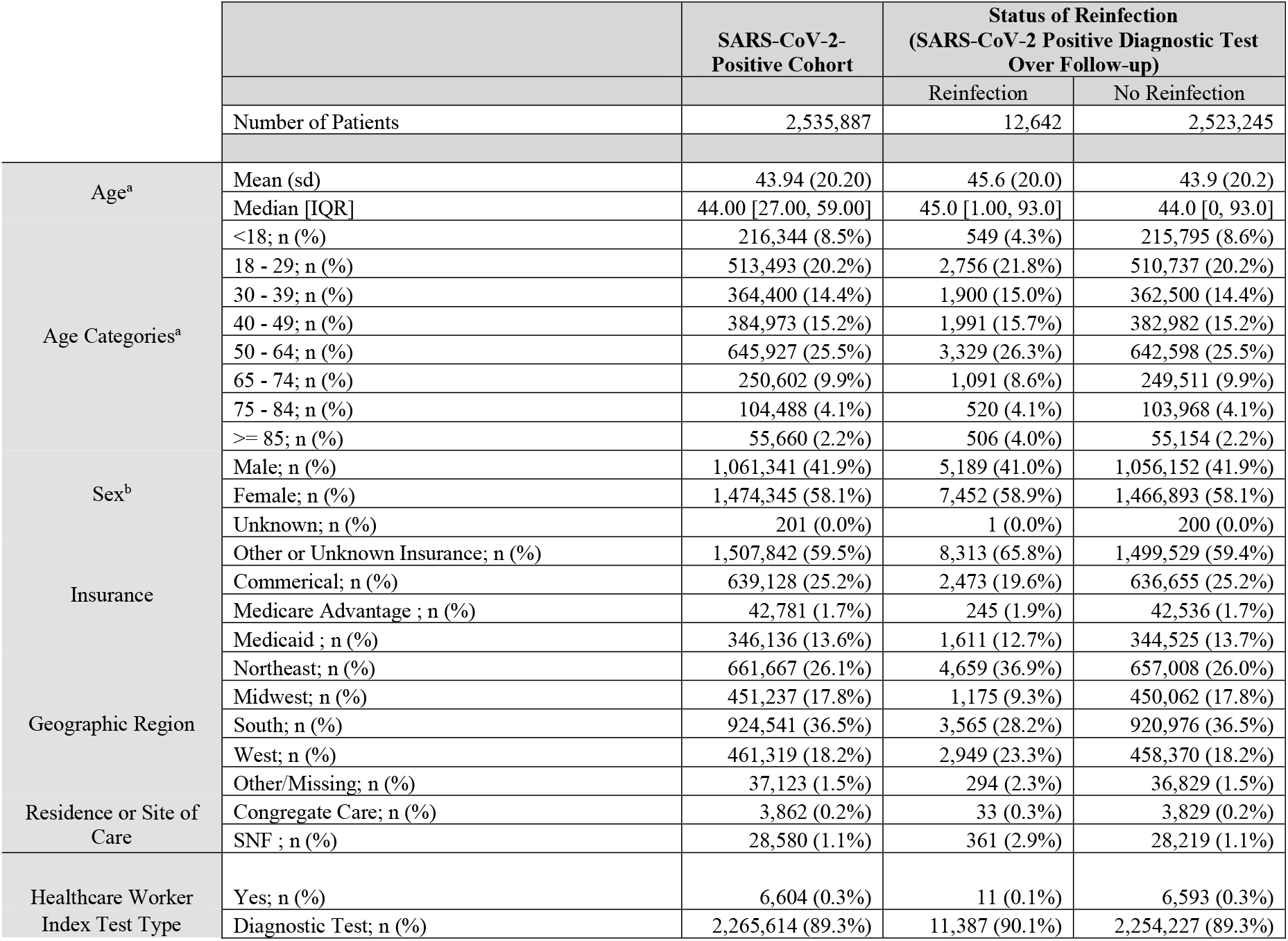
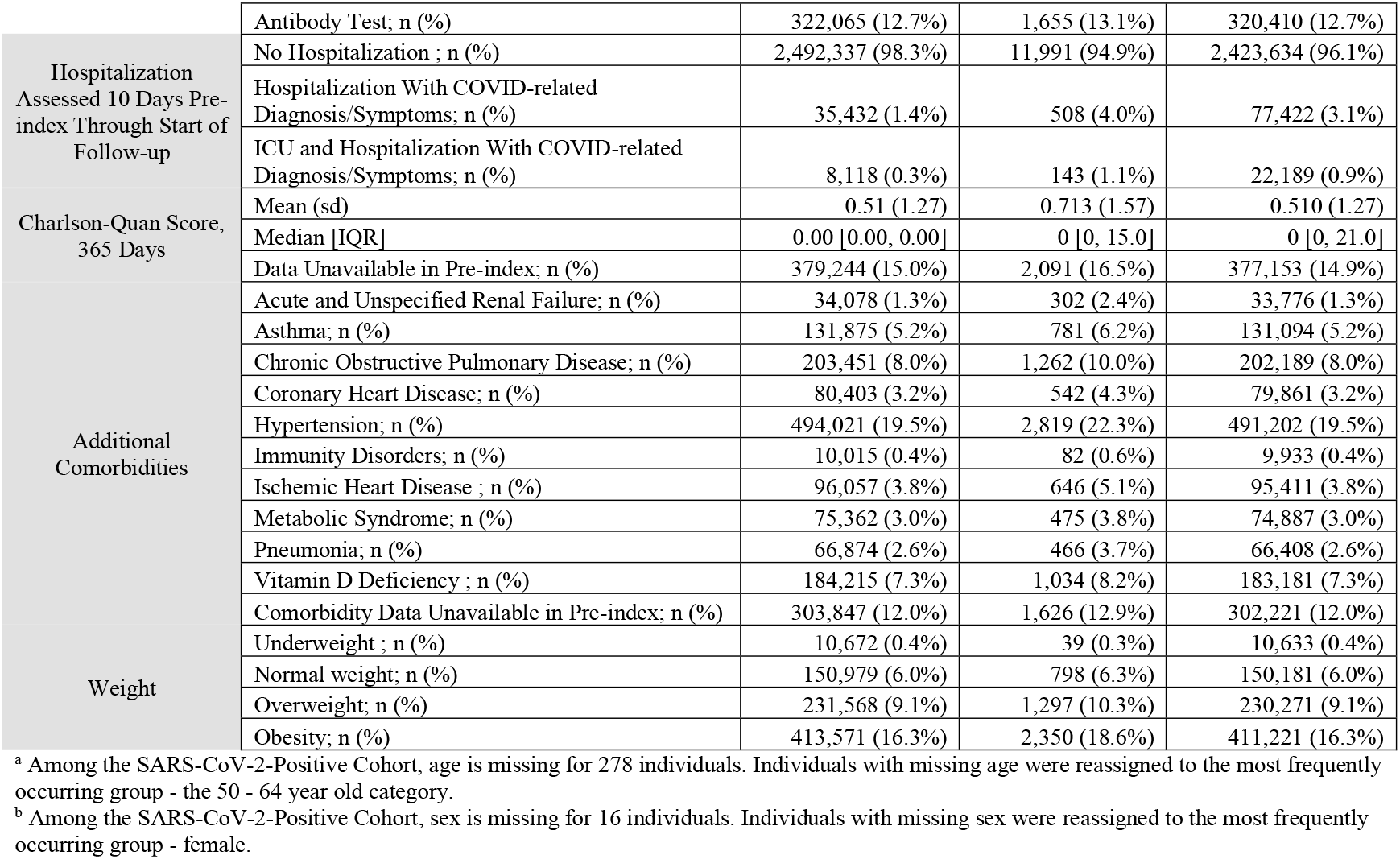
Demographics and baseline comorbidities in the 365 days prior to first SARS-CoV-2 positive test among a SARS-CoV-2-positive cohort identified between February 29th, 2020 through December 9th, 2020

During follow-up, the crude rate of reinfection was 11.75 (CI: 11.55, 11.96) per 1,000 PY; 12,642 cases over 1,075,563 PY. Individuals ≥85 years of age were more likely to be reinfected compared to those aged 18-29 years. This was particularly true in the Midwest and South regions. Additionally, individuals living in a skilled nursing facility (SNF) or in congregate care settings were 1.5 and 2.8 times as likely to be reinfected compared to those not residing in these settings on the index date, respectively. In the South and Midwest geographies, those in congregate care settings had a 4-times higher risk compared to those not living in congregate care settings. Patients with comorbid immunologic conditions had a 37% higher risk of being reinfected than those without. The risk of reinfection among other comorbid conditions, such as heart failure, varied by geographic region. Healthcare workers were less than half as likely to be reinfected compared to the general population. (Table 4). These results suggest that immunity to SARS-CoV-2 reinfection may be less durable in elderly patients, those living in congregate setting, and in individuals with impaired immune function.

**Table 4.**
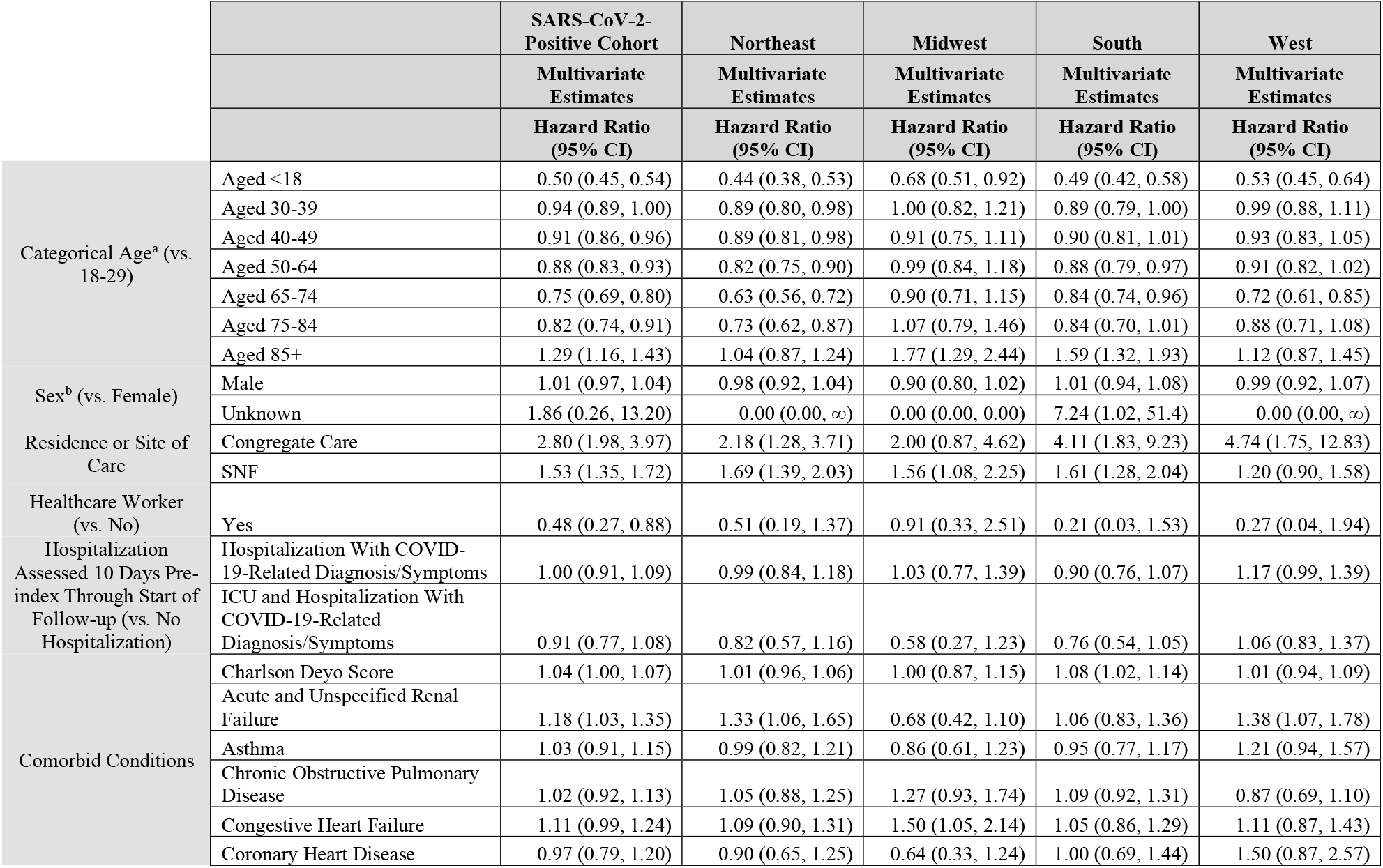
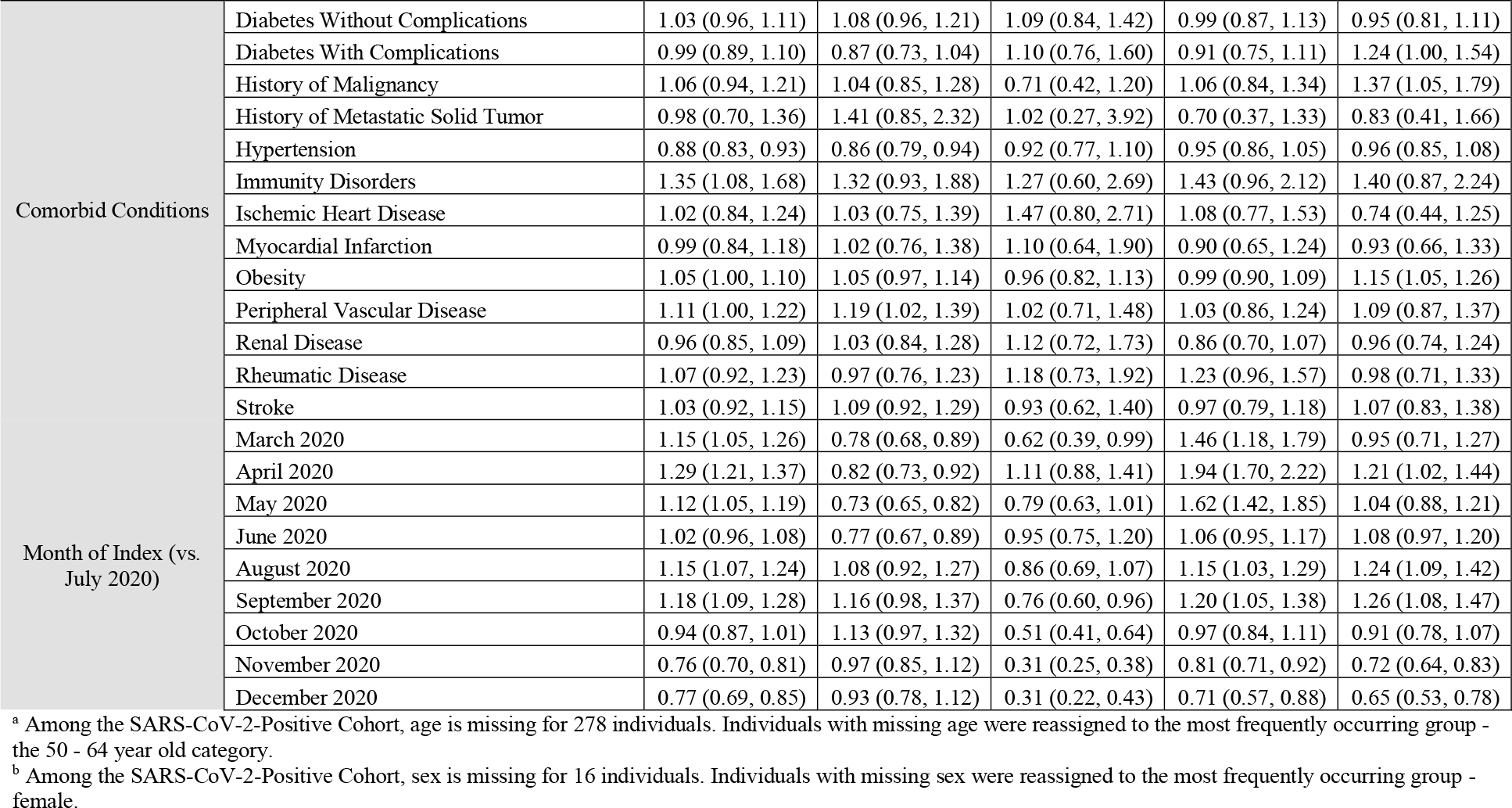
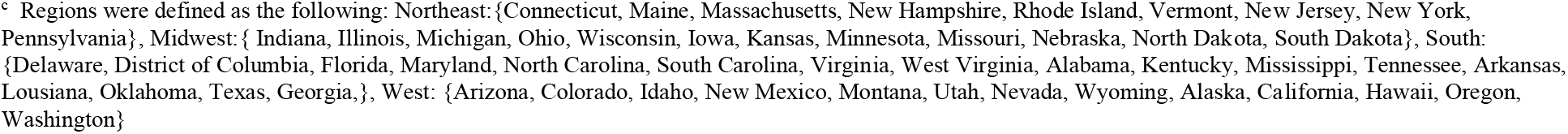
Measured association of demographic and clinical factors with re-infection (SARS-CoV-2 positive diagnostic test during follow-up (April 30th, 2020 through April 30th, 2021)) among individuals with history of SARS-CoV-2 infection identified between February 29th, 2020 through December 9th, 2020

## Discussion

Among a cohort of >22 million individuals with SARS-CoV-2 laboratory test results, the risk of reinfection in index-positive individuals was 87% lower compared to risk of infection among index-negative individuals. This association was consistent in unadjusted, fully-adjusted, and among propensity-score matched model estimates. The observed protection against reinfection was durable for at least one year. Factors associated with an increased likelihood of reinfection included older age, comorbid immunological conditions, and living in congregate care setting. Consistent with other studies, healthcare workers were half as likely to be reinfected compared to the general population, which may be associated with lifestyle factors and/or activities to reduce transmission, such as social distancing and use of mask wearing.^18–20^

Our prior work suggested 90% protection in a real-world cohort of 3.2 million individuals.^4^ In this current analysis, the observed magnitude of the decreased risk of infection is consistent with other limited published estimates that have been obtained with smaller cohorts that mainly examined shorter periods of protection.^15, 19, 21^ This work adds to our prior findings showing that the real-world use of widely available diagnostics (antibody assays and NAAT) can identify individuals with prior infection and reliably predict long-term risk of reinfection.

Consistent with findings from this study, in a recent US study among over 325,000 patients from a health system spanning two states who were PCR tested for SARS-CoV-2 between March 2020 and September 2021, the duration of long-term protection afforded from primary SARS-CoV-2 infection was up to 13 months.^22^ A population-based study of over 500,000 individuals in Denmark evaluating infection measured by PCR testing during the second COVID-19 surge among patients who were tested in the first COVID-19 surge, suggest duration lasting at least 7 months, as no waning immunity was observed when comparing results at 3-6 months vs. ≥7 months.^21^ Further, a meta-analysis conducted in 2021 suggested that immunity from primary SARS-COV-2 infection likely persists through one year.^23^ This present study now extends those observations by examining a much larger, real-world population followed for over a year.

The stable level of protection from reinfection observed in this study through the first year after SARS-CoV-2 infection differs from the duration of protection after two doses of a SARS-CoV-2 mRNA vaccine, which has been reported to decrease after a few months.^24^ This difference may not be surprising, given what is known about duration of protection from other subunit vaccines compared with duration of protection following viral infection^25^, and the much more rapid reported decrease in antibody titer following two doses of a SARS-CoV-2 mRNA vaccine compared with the rate of decrease after viral infection.^26^

Of importance to future studies utilizing real-world evidence and diagnostic codes where more reliable laboratory data aren’t readily available as in our study, when ICD-10 diagnostic codes were added to the primary outcome definition, we found a notably lower risk reduction (27%) for reinfection vs. infection. This suggests the lack of specificity in COVID-19 ICD-10 codes. Other studies have found variable positive predictive value of COVID ICD-10 codes based on care setting (i.e., inpatient or outpatient).^27–30^

The present study identified a large national population using data from medical and pharmacy claims, retail pharmacy data, and electronic medical records. This large size and broad representation across many evaluated attributes allowed better characterization of subgroups. These data are not specifically intended for research purposes; thus, the completeness of medical information is unknown. Additionally, certain risk factors for infection, such as frequency of exposure to SARS-CoV-2 are not captured in the RWD. We were, therefore, not able to assess social and behavioral factors that likely influence risk of reinfection, which may be why we observe the same effect estimates in the crude and adjusted models

These data are mainly drawn from a medically insured individuals, except for laboratory or retail pharmacy data coming directly from available clinical laboratory and retail pharmacy sources. As such, these data may not be representative of the medically uninsured individuals, although there is no a priori reason to believe their results would be different. During follow-up, index negative individuals had slightly more follow-up time and NAAT tests than index-positive, which may overestimate the true protective effect observed. Further, false-negative NAAT test results among the index-negative may underestimate the true protective effect observed.^31^. Study data aren’t inclusive of time periods of variant circulation (i.e. Delta, Omicron); durability of protection may vary for these and future SARS-CoV-2 variants.

The primary outcome definition inherently required individuals to be observable, and without evidence of positive test results if in the index-negative group, until 61 days following their index date, thereby introducing immortal time bias (i.e. bias in the estimator due to exclusion of time intervals, in this case 60 days post-index). However, this decision was warranted to ensure residual viral shedding wasn’t captured during a primary infection, creating a more specific outcome definition. ^9–11^

In summary, this large US population-based study demonstrates that SARS-CoV-2 reinfection is uncommon among individuals with laboratory evidence of a previous infection. Protection from SARS-CoV-2 reinfection is stable for up to one year. Reinfection risk was primarily associated with age 85+ years, comorbid immunologic conditions and living in congregate care settings; healthcare workers demonstrated a decreased reinfection risk. These findings suggest that infection induced immunity is durable for variants circulating prior to Delta.

## Supporting information

Supplemental eMethods

## Data Availability

All data produced in the present study are available upon reasonable request to the authors.

## Acknowledgement

All authors completed the ICMJE uniform disclosure form at: www.icmje.org/coi_disclosure.pdf and declare: Shannon L. Reynolds and Christopher Bush had full access to all the data and take full responsibility for the integrity of the data and the accuracy of the data analysis. Shannon L. Reynolds, Carly Kabelac, and Christopher Bush are employees of and own stock in Aetion, Inc. Harvey W. Kaufman and William A. Meyer III are employees of and own stock in Quest Diagnostics. Oren Cohon and Steve Anderson are employees of and own stock in Labcorp Drug Development. Steve Anderson has received consulting fees from Luminex. Douglas Lowry has received royalty-related payments from NIH. Kathy Cronin, Sandy Leonard, Valentina Petkov, Norman Sharpless, and Lynne Penberthy report no conflict of interests. The authors would like to acknowledge Wendy Turenne for her support on this engagement and Reyna Klesh for her data expertise support.

