## Supplemental eMethods for "Duration of Protection Against SARS-CoV-2 Reinfection and Associated Risk of Reinfection Assessed with Real-World Data"

| **Supplementary Methods** | |
| --- | --- |
| eMethods1 | Pre-exposure covariates used to describe baseline characteristics and for confounding adjustment via propensity score matching |
| eMethods2 | Codes of pre-exposure covariates used to describe baseline characteristics and for confounding adjustment via propensity score matching |

### **Supplementary Methods**

### **eMethods1: Pre-exposure covariates used to describe baseline characteristics and for confounding adjustment via propensity score matching**

Variables listed were included as baseline characteristics and model covariates. Presence of comorbidities was determined based on whether they ever occurred during the 12-month baseline period prior to the index date, unless specified otherwise, while demographic and SARS-CoV-2 test characteristics were assessed on the index date unless otherwise noted. The symbol * denotes variables included in the propensity-score model.

**Variable name:**

Month/Year of Index*, Age* on index, Categorical Age on index, Gender* on index, Insurance Category* on index, U.S. Region* on index, State on index, Congregate Care* on index, SNF* on index , Healthcare Worker* on index, Diagnostic Test* on index, Antibody Test* on index, Charlson-Quan score over prior 365 days*, HIV/AIDS, CHF, COPD, Dementia, Diabetes without complications, Diabetes with complications, Hemiplegia or Paraplegia, Malignancy, Metastatic Solid Tumor, MI, Mild Liver Disease, Moderate or Severe Liver Disease, Peptic Ulcer Disease, Peripheral Vascular Disease, Renal Disease, Rheumatic Disease, Stroke, Pneumonia*, Asthma*, Chronic Obstructive Pulmonary Disease (COPD)*, Immunity Disorders*, Acute and unspecified renal failure*, Ischemic heart disease*, Hypertension*, Coronary Heart Disease*, Metabolic Syndrome*, Vitamin D Deficiency*, Underweight*, Normal weight*, Overweight*, Obesity*, Patients with Available Claims or Charge Master Data Pre-index, Patients with Available Claims or Chargemaster or EHR Data Pre-index

### **eMethods2: Codes of pre-exposure covariates used to describe baseline characteristics and for confounding adjustment via propensity score matching**

The symbol * denotes variables included in the propensity-score model.

| **Variable** | **Definition** |
| --- | --- |
| Demographic Characteristics | |
| Age* | Continuous numeric variable |
| Age, categorical | Age is between 0-4 (ref), 5-10, 11-15, 16-17, 18-29, 30-39, 40-49, 50-64, 65-74, 75-84, 85 or more |
| Sex* | Male/Female/Unknown/Missing |
| Insurance category* | Missing/Commercial/Medicaid/Medicare |
| U.S region* | Northeast: Connecticut, Massachusetts, Vermont, Rhode Island, Maine, New York, New Jersey, New Hampshire, Pennsylvania  Midwest: Iowa, Illinois, Indiana, Kansas, Michigan, Minnesota, Missouri, North Dakota, Nebraska, Ohio, South Dakota, Wisconsin  South: Alabama, Arkansas, District of Columbia, Delaware, Florida, Georgia, Kentucky, Louisiana, Maryland, Mississippi, North Carolina, Oklahoma, South Carolina, Tennessee, Texas, Virginia, West Virginia  West: Alaska, Arizona, California, Colorado, Hawaii, Idaho, Montana, New Mexico, Nevada, Oregon, Utah, Washington, Wyoming  Other/Missing/Unknown: Armed Forces Americas, Armed Forces, Armed Forces Pacific, American Samoa, Micronesia, Guam, Marshall Islands, Northern Marianas Islands, Puerto Rico, Palau, Virgin Islands, Missing |
| Congregate care* | Living in Congregate Care Indicator is Yes. Sourced from laboratory data |
| Skilled nursing facility^1-2^* | Medical Claim with any of the following place of service, HCPCS/CPT, bill code or revenue code:  Place of Service: “Skilled Nursing Facility” or “Nursing Facility”  HCPCS/CPT code: 99303, 99307, 99308, 99310, 99311, 99312, 99313, 99315, 99316, 94004, 99301, 99302, 99304, 99305, 99306, 99309, 99318  Type of Bill Code: 2*  Revenue Code: 0552, 0553, 0556, 0022, 0550, 0551, 0559  Hospital Chargemaster Data with any of the following department code, HCPCS/CPT, admission source or discharge status:  Department Codes: 0550, 10088000, 3159, 10083900, 37230, 5023  HCPCS/CPT code of 99303, 99307, 99308, 99310, 99311, 99312, 99313, 99315, 99316, 94004, 99301, 99302, 99304, 99305, 99306, 99309, 99318  Admission Source: Transferred from a Skilled Nursing Facility  Discharge Status: Discharged/Transferred to a Nursing Facility Cert Under Medicare |
| Healthcare worker status* | Employed in Healthcare Indicator is Yes. Sourced from laboratory data. |
| Month/Year of index date* | The month and year that an individual indexes into the cohort. |
| Available claims or chargemaster data in the year prior to index | Presence of a claim or chargemaster data in the 12 months prior to the index date. |
| Available claims or EHR or chargemaster data in the year prior to index | Presence of a claim or EHR or chargemaster data in the 12 months prior to the index date. |
| Index Test Characteristics | |
| Diagnostic test result* | Record of any positive or negative result from a COVID-19 diagnostic laboratory test. |
| Antibody test result* | Record of any positive or negative result from a COVID-19 antibody laboratory test. |
| Vaccine Manufacturer Definitions^3^  *Note: CVX codes indicate the product used in vaccination and are maintained by the CDC’s National Center of Immunization and Respiratory Diseases.* | |
| Moderna | Moderna vaccines were defined using the Medical or Pharmacy Claims with the following NDC or Procedure Codes:  NDC: 08077702371, 08777027399, 80770027399, 80777023799, 80777027309, 80777027310, 80777027398, 80777027699, 80777273990, 08077727310, 08077727399, 80777027399, 80777072310, 80777073100, 80777273099, 8077727310, 8077727399, 80777727310, 87770273980  Procedure code: 0011A, 0012A, 91301  Or Retail Pharmacy Data with the following Manufacturer: “MOD,” “MODERNA,” “MODERNA US, INC,” “MODERNA US, INC.,”  Category Code Type “CVX_CODE” with Code ‘207’ |
| Pfizer | Pfizer Vaccines were defined using the Medical or Pharmacy Claims with the following NDC or Procedure Codes:  NDC: 52967100002, 59262100002, 59267010001, 592671000002, 59267100001, 59267100002, 59267100003, 59267100010, 05926710001, 58267100001, 59267010002, 5926710001, 59267100020  Procedure code: 0001A, 0002A, 91300  Or Retail Pharmacy Data with the following Manufacturer: “PFIZER,” “PFIZER, INC,” “PFR”  Category Code Type “CVX_CODE” with Code ‘208’ |
| J&J | J&J Vaccines were defined using the Medical or Pharmacy Claims with the following NDC or Procedure Codes:  NDC: 5967658015, 05967658005, 59676000000, 59676058000, 59676058005, 59676058015, 59676059005, 59676580015, 5967658005, 59676580150  Procedure codes: 0031A, 91303  Or Retail Pharmacy Data with the following Manufacturer: “JANSSEN,” “JANSSEN (DIVISION OF JOHNSON AND JOHNSON),” “JOHNSON AND JOHNSON,” “JSN”  Category Code Type “CVX_CODE” with Code ‘212’ |
| COVID-19 Severity (Assessed only for *SARS-CoV-2-Positive Cohort* from 10 days prior to index to day 60) | |
| No Hospitalization with COVID-19 diagnosis or related symptom* | Neither hospitalized with a COVID-19 diagnosis or symptom nor hospitalized in the ICU with a COVID-19 diagnosis or symptom |
| Hospitalized with a COVID-19 diagnosis or related symptom* | Medical or Chargemaster Data indicating a hospitalization with one of the following ICD-10 diagnosis codes:   - B97.29 - Other coronavirus as the cause of diseases classified elsewhere - J02.9 - Acute pharyngitis, unspecified - J12.81 - Pneumonia due to SARS-associated coronavirus - J18.1 - Lobar pneumonia, unspecified organism - J18.8 - Other pneumonia, unspecified organism - J20.8 - Acute bronchitis due to other specified organisms - J40 - Bronchitis, not specified as acute or chronic - J80 - Acute respiratory distress syndrome - J85.1 - Abscess of lung with pneumonia - J96.0 - Acute respiratory failure - J96.01 - Acute respiratory failure with hypoxia - J96.02 - Acute respiratory failure with hypercapnia - J96.2 - Acute and chronic respiratory failure - J96.21 - Acute and chronic respiratory failure with hypoxia - J98.8 - Other specified respiratory disorders - K59.1 - Functional diarrhea - R04.2 - Hemoptysis - R06.0 - Dyspnea - R06.00 - Dyspnea, unspecified - R06.01 - Orthopnea - R06.2 - Wheezing - R07.8 - Other chest pain - R07.89 - Other chest pain - R09.2 - Respiratory arrest - R11.2 - Nausea with vomiting, unspecified - R43.1 - Parosmia - R43.2 - Parageusia - R43.8 - Other disturbances of smell and taste - R43.9 - Unspecified disturbances of smell and taste - R50.9 - Fever, unspecified - R51 - Headache - R68.83 - Chills (without fever) - U07.2 - COVID-19, virus not identified - Z20.828 - Contact with and (suspected) exposure to other viral communicable diseases - B34.2 - Coronavirus infection, unspecified - B34.9 - Viral infection, unspecified - B97.21 - SARS-associated coronavirus as the cause of diseases classified elsewhere - J06.9 - Acute upper respiratory infection, unspecified - J12.89 - Other viral pneumonia - J12.9 - Viral pneumonia, unspecified - J18.0 - Bronchopneumonia, unspecified organism - J18.9 - Pneumonia, unspecified organism - J22 - Unspecified acute lower respiratory infection - J96.00 - Acute respiratory failure, unspecified whether with hypoxia or hypercapnia - J96.20 - Acute and chronic respiratory failure, unspecified whether with hypoxia or hypercapnia - J96.22 - Acute and chronic respiratory failure with hypercapnia - J96.90 - Respiratory failure, unspecified, unspecified whether with hypoxia or hypercapnia - J98.4 - Other disorders of lung - M79.1 - Myalgia - M79.10 - Myalgia, unspecified site - M79.11 - Myalgia of mastication muscle - M79.12 - Myalgia of auxiliary muscles, head and neck - M79.18 - Myalgia, other site - R05 - Cough - R06.02 - Shortness of breath - R06.03 - Acute respiratory distress - R06.09 - Other forms of dyspnea - R07.1 - Chest pain on breathing - R07.9 - Chest pain, unspecified - R09.02 - Hypoxemia - R09.3 - Abnormal sputum - R11.0 - Nausea - R11.10 - Vomiting, unspecified - R11.11 - Vomiting without nausea - R11.12 - Projectile vomiting - R19.7 - Diarrhea, unspecified - R43 - Disturbances of smell and taste - R43.0 - Anosmia - R91.8 - Other nonspecific abnormal finding of lung field - U07.1 - COVID-19 |
| Hospitalized in the ICU with a COVID-19 * | Hospitalized with a COVID-19 diagnosis or related symptom (defined above) with a medical claim revenue code or chargemaster department code indicating an ICU hospitalization. |
| Charlson-Quan Comorbidities^4^ | |
| Charlson-Quan Comorbidity Index* | Composite score based on the below components, assessed in the 365 days prior to index date |
| AIDS component | ICD-10: B20.x–B22.x, B24.x |
| CHF component | ICD-10: I09.9, I11.0, I13.0, I13.2, I25.5, I42.0, I42.5–I42.9, I43.x, I50.x, P29.0 |
| COPD component | ICD-10: I27.8, I27.9, J40.x–J47.x, J60.x–J67.x, J68.4, J70.1, J70.3 |
| Dementia component | ICD-10: F00.x–F03.x, F05.1, G30.x, G31.1 |
| Hemiplegia or Paraplegia component | ICD-10: G04.1, G11.4, G80.1, G80.2, G81.x, G82.x, G83.0–G83.4, G83.9 |
| Malignancy component | ICD-10: C00.x–C26.x, C30.x–C34.x, C37.x–C41.x, C43.x, C45.x–C58.x, C60.x–C76.x, C81.x–C85.x, C88.x, C90.x–C97.x |
| Metastatic Solid Tumor component | ICD-10:C77.x–C80.x |
| MI component | ICD-10: I21.x, I22.x, I25.2 |
| Diabetes without Chronic Complications component | ICD-10: E10.0, E10.1, E10.6, E10.8, E10.9, E11.0, E11.1, E11.6, E11.8, E11.9, E12.0, E12.1, E12.6, E12.8, E12.9, E13.0, E13.1, E13.6, E13.8, E13.9, E14.0, E14.1, E14.6, E14.8, E14.9 |
| Mild Liver Disease component | ICD-10: B18.x, K70.0–K70.3, K70.9, K71.3–K71.5, K71.7, K73.x, K74.x, K76.0, K76.2–K76.4, K76.8, K76.9, Z94.4 |
| Moderate or Severe Liver Disease component | ICD-10: I85.0, I85.9, I86.4, I98.2, K70.4, K71.1, K72.1, K72.9, K76.5, K76.6, K76.7 |
| Peptic Ulcer Disease component | ICD-10: K25.x–K28.x |
| Peripheral Vascular Disease component | ICD-10: I70.x, I71.x, I73.1, I73.8, I73.9, I77.1, I79.0, I79.2, K55.1, K55.8, K55.9, Z95.8, Z95.9 |
| Renal Disease component | ICD-10: I12.0, I13.1, N03.2–N03.7, N05.2–N05.7, N18.x, N19.x, N25.0, Z49.0–Z49.2, Z94.0, Z99.2 |
| Rheumatologic Disease component | ICD-10: M05.x, M06.x, M31.5, M32.x–M34.x, M35.1, M35.3, M36.0 |
| Diabetes with Chronic Complications component | ICD-10: E10.2–E10.5, E10.7, E11.2–E11.5, E11.7, E12.2–E12.5, E12.7, E13.2–E13.5, E13.7, E14.2–E14.5, E14.7 |
| Cerebrovascular Disease component | ICD-10: G45.x, G46.x, H34.0, I60.x–I69.x |
| Additional Comorbidities | |
| Pneumonia* | ICD-10: J12.9, J85.1 J18.8, J18.0, J12.81, J12.89, J18.1, J18.9 |
| Asthma^6^* | HCUP CCSR RSP009: J45.21, J45.31, J45.50, J45.20, J45.30, J45.32, J45.41, J45.902,  J45.998, J45.51, J45.901, J45.990, J45.991, J45.22, J45.40, J45.42,  J45.52, J45.909 |
| Chronic obstructive pulmonary disease (COPD)^5^* | HCUP CCSR RSP008: J41.0, J41.1, J41.8, J42, J43.8, J43.9, J44.0, J44.9, J47.1, J43.1,  J47.0, J43.0, J47.9, J43.2, J44.1 |
| Acute and unspecified renal failure (Renal impairment)^5^* | HCUP CCSR GEN002: N99.0, N17.1, N19, O90.4, N17.9, N17.0, N17.2, N17.8 |
| Ischemic Heart Disease^6^* | ICD-10: I20, I23, I24, I25 |
| Immunity Disorders^5^* | HCUP CCSR BLD008: D80.3, D80.4, D81.89, D83.1, D86.1, D86.2, D89.40, D89.41, D89.43, D89.811, D80.1, D80.6, D81.1, D81.2, D81.6, D81.810, D81.818, D82.0, D82.2, D82.3, D82.9, D83.0, D83.8, D84.1, D86.82, D86.83, D86.84, D86.85, D89.2, D89.812, D80.2, D81.0, D81.7, D81.819, D82.8, D83.2, D83.9, D84.8, D86.87, D86.89, D86.9, D89.0, D89.1, D89.82, D89.9, D80.5, D80.8, D81.4, D82.4, D86.0, D86.86, D89.42, D89.813, D80.0, D80.7, D80.9, D81.3, D81.5, D81.9, D82.1, D84.0, D84.9, D86.3, D86.81, D89.3, D89.49, D89.810, D89.89 |
| Hypertension* | ICD-10: I10, I11, I12, I13, I15, I16  Investigator derived |
| Coronary Heart Disease* | ICD-10:I25.10  Investigator derived |
| Metabolic Syndrome^7^* | Three or more of the five below  High fasting glucose ICD-10 [R73]  High blood pressure/ hypertension ICD-10 [R03, I10]  Lipoprotein deficiency ICD-10 [E78.6]  Hypertriglyceridemia ICD-10 [E78.1]  Obesity ICD-10 [Z68.30 to Z68.45, E66, O99.2] or BMI > 30 (weight (kg) / [height (m)]2) |
| Vitamin D Deficiency* | E55. 9  Investigator derived |
| Underweight^8^* | ICD-10: Z68.1 and R63.6, or R63.6  EMR-based: BMI < 18.5 (weight (kg) / [height (m)]2) |
| Normal Weight* | ICD-10: Z68.1 and no R63.6, or Z68.20 - Z68.24  EMR-based: BMI 18.5 < 24.9 (weight (kg) / [height (m)]2)  Investigator derived |
| Overweight* | ICD-10: Z68.25 to Z68.29, E66.3  EMR-based: BMI 25.0 < 29.9 (weight (kg) / [height (m)]2)  Investigator derived |
| Obese* | ICD-10: Z68.30 to Z68.45, E66, O99.2  EMR-based: BMI > 30 (weight (kg) / [height (m)]2)  Investigator derived |
